# Reductions in US life expectancy during the COVID-19 pandemic by race and ethnicity: Is 2021 a repetition of 2020?

**DOI:** 10.1101/2021.10.17.21265117

**Authors:** Theresa Andrasfay, Noreen Goldman

## Abstract

COVID-19 had a huge mortality impact in the US in 2020 and accounted for most of the overall reduction in 2020 life expectancy at birth. There were also extensive racial/ethnic disparities in the mortality impact of COVID-19 in 2020, with the Black and Latino populations experiencing reductions in life expectancy at birth over twice as large as that of the White population. Despite continued vulnerability of these populations, the hope was that widespread distribution of effective vaccines would mitigate the overall impact and reduce racial/ethnic disparities in 2021. In this study, we quantify the mortality impact of the COVID-19 pandemic on 2021 US period life expectancy by race and ethnicity and compare these impacts to those estimated for 2020. Our estimates indicate that racial/ethnic disparities have persisted, and that the US population experienced a decline in life expectancy at birth in 2021 of 2.2 years from 2019, 0.6 years more than estimated for 2020. The corresponding reductions estimated for the Black and Latino populations are slightly below twice that for Whites, suggesting smaller disparities than those in 2020. However, all groups experienced additional reductions in life expectancy relative to 2020, and this apparent narrowing of disparities is primarily the result of Whites experiencing proportionately greater increases in mortality in 2021 compared with the corresponding increases in mortality for the Black and Latino populations in 2021. Estimated declines in life expectancy at age 65 increased slightly for Whites between 2020 and 2021 but decreased for both the Black and Latino populations, resulting in the same overall reduction (0.8 years) estimated for 2020 and 2021.

## Introduction

The staggering death toll in the US from COVID-19 has been well-documented: deaths attributed to COVID-19 in 2020 account for most of the 1.8-year reduction in period life expectancy at birth, reversing over 18 years of progress in mortality improvement (1,2). In a previous paper, we predicted that widespread availability of an effective vaccine would lessen the impact of COVID-19 on 2021 life expectancy compared with 2020, although we argued that life expectancy was unlikely to return to pre-pandemic levels (3). Several highly effective vaccines have indeed been developed in record time, but relatively low vaccine coverage in the US, combined with the highly transmissible Delta and Omicron variants of SARS-CoV-2, have led to additional mortality surges, and, by the end of 2021, the total number of COVID-19 deaths had exceeded the 2020 total by 30% (4–6). These sobering numbers combined with the younger age distribution of deaths in 2021 (7) (see Fig 1) – resulting partly from higher vaccination rates among older individuals – indicate that life expectancy in 2021 may be even lower than that in the preceding year.

**Fig 1.**
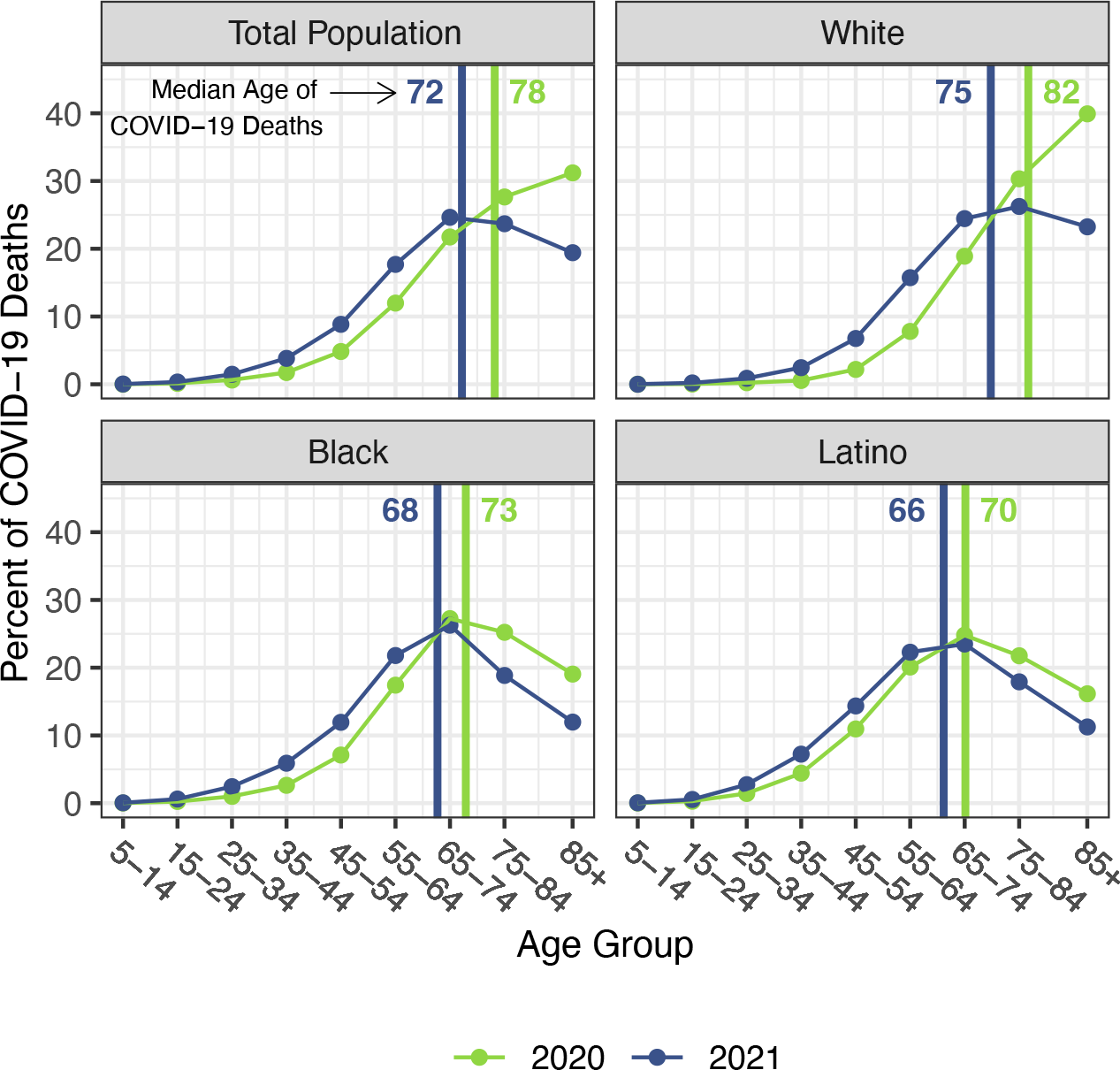
Percentage of COVID-19 deaths in each age group by race/ethnicity, 2020 and 2021. Vertical lines indicate the median age of COVID-19 deaths in each year. We estimate the median age of COVID-19 deaths in both years with the assumption that deaths occur uniformly within each age group. Data are from CDC WONDER (July 6, 2022, update). Deaths below age 5 are not shown because some counts are suppressed.

The disproportionate impact of COVID-19 on the survival of vulnerable populations has also received extensive attention: the Latino and Black populations experienced declines in life expectancy over twice as large as that for Whites in 2020 (1). Risk factors for COVID-19 infection and mortality, such as crowded living conditions, frontline jobs with high exposure to infection and low pay, dependence on public transportation, low access to quality healthcare, and high rates of select chronic conditions, still characterize these groups, suggesting continued racial/ethnic disparities in COVID-19 mortality (8–12). A strategically targeted vaccine distribution had the potential to reduce racial/ethnic disparities in COVID-19 mortality in 2021 (13), but many individuals faced barriers to vaccination in the early months, including difficulty scheduling vaccine appointments online, limited supplies, lack of transportation to vaccination sites, and lack of time off work to get vaccinated and recover from side effects (14,15). The resulting inequitable vaccine distribution and uptake may have further exacerbated racial/ethnic disparities in COVID-19 mortality. However, as vaccines became more widely available in the later months of 2021, racial/ethnic differentials in vaccination rates decreased (16,17). By August 2021, differences in vaccination rates by political affiliation, religion, and rural/urban status exceeded racial/ethnic differences (16), suggesting a potential reduction of racial/ethnic disparities in COVID-19 mortality in 2021 relative to those in 2020.

Our goal in the current study is to estimate racial and ethnic disparities in the mortality impact of the COVID-19 pandemic in 2021 and establish the extent to which these disparities differed from those in the previous year. We do so by calculating reductions in period life expectancy at birth and at age 65 relative to the pre-pandemic levels for each of these populations. Such period life expectancy reductions are a useful metric during a pandemic because they capture the mortality impact of the pandemic in a single measure that can be compared across subgroups, nations, or years. We incorporate deaths from all causes in these calculations because it has been well-established that the pandemic has resulted in increases in numerous causes of death other than COVID-19 (1,18,19).

## Data and Methods

### Data

Age-specific mortality rates of deaths from all causes and deaths for which COVID-19 was the underlying cause are obtained for 2020 and 2021 for each race/ethnicity from the CDC WONDER (Wide-ranging Online Data for Epidemiologic Research) database as of July 7, 2022 (CDC WONDER 2022). CDC WONDER provides publicly available and user-friendly mortality, population, and other public health data. Deaths for 2020 are considered final and complete, but deaths for 2021 are still provisional and may therefore be subject to reporting delays. CDC WONDER data for both 2020 and 2021 use the same July 1, 2020 population counts that are projected from the 2010 Census as denominators for mortality rates. We adjust the race/ethnicity-specific mortality rates in 2020 and 2021 to account for racial misclassification by multiplying mortality rates by the age-group and race-specific classification ratios published by NVSS. These ratios use a linkage of individuals in the Current Population Survey (CPS) or the census to death certificates for deaths between 1999-2011 to estimate the degree of misclassification of race/ethnicity recorded on death certificates compared with self-reported information in the CPS or census (20). We use these classification ratios with the assumption that misclassification in 2020 and 2021 for all causes of death, including COVID-19, is the same magnitude as in these previous years.

## Methods

Before calculating the life tables for 2020 and 2021, we present the following three sets of statistics specific to COVID-19 deaths to understand how racial/ethnic disparities in COVID-19 mortality have changed over the two years: the age distribution of COVID-19 deaths, age-specific COVID-19 mortality rates, and the ratios of COVID-19 mortality rates for the Black and Latino populations relative to Whites. Next, for each population, we create life tables for 2020 and 2021 that incorporate mortality from all causes of death. We borrow values of person-years lived by infants dying before age 1 (1a0) from the 2019 life tables published by NVSS (20). We use the mortality rates adjusted for racial misclassification and standard life table relationships to calculate the remaining life table columns and obtain estimates of life expectancy at each age (21). These life tables are estimated with single-year age-intervals up to age 85 and an open-ended age interval for 85 and older.

## Results

Figs 1-3 document changing patterns in COVID-19 deaths between 2020 and 2021. Fig 1, which presents the percentage of COVID-19 deaths in each age group and the approximate median age of COVID-19 deaths in each year, reveals that for all populations, but especially for the White population, the distribution of COVID-19 deaths has shifted to younger ages. Fig 2, which presents age-specific COVID-19 death rates for each of the past two years, portrays a similar time pattern across racial/ethnic groups: a large decline in rates at the oldest ages between 2020 and 2021, with modest changes – often small increases – below age 65. Fig 3, which shows these rates for the Black and Latino populations divided by the corresponding rates for the White population, highlights the huge decline in relative death rates for the working age population (18-65): COVID-19 death rates at these ages were often three to five times as high in the Black and Latino populations as among Whites in 2020 but roughly twice as high in 2021. These numbers suggest a substantial decline in racial/ethnic disparities in COVID-19 mortality from 2020 to 2021, but the estimates of life expectancy provided below tell a more nuanced story.

**Fig 2.**
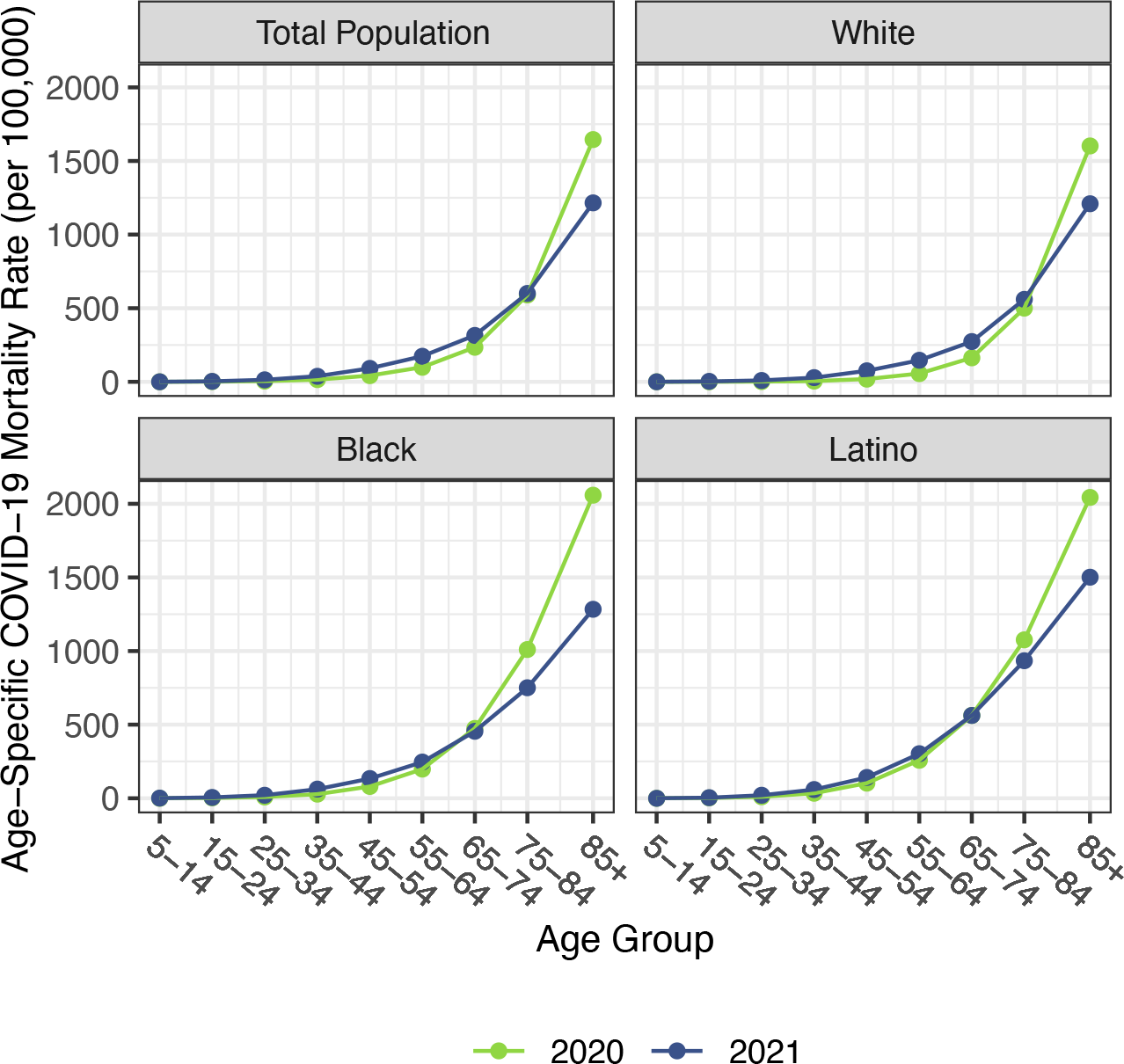
Age-specific COVID-19 mortality rates by race/ethnicity, 2020 and 2021. Data are from CDC WONDER (July 6, 2022, update). Mortality rates below age 5 are not shown because some counts are suppressed.

**Fig 3.**
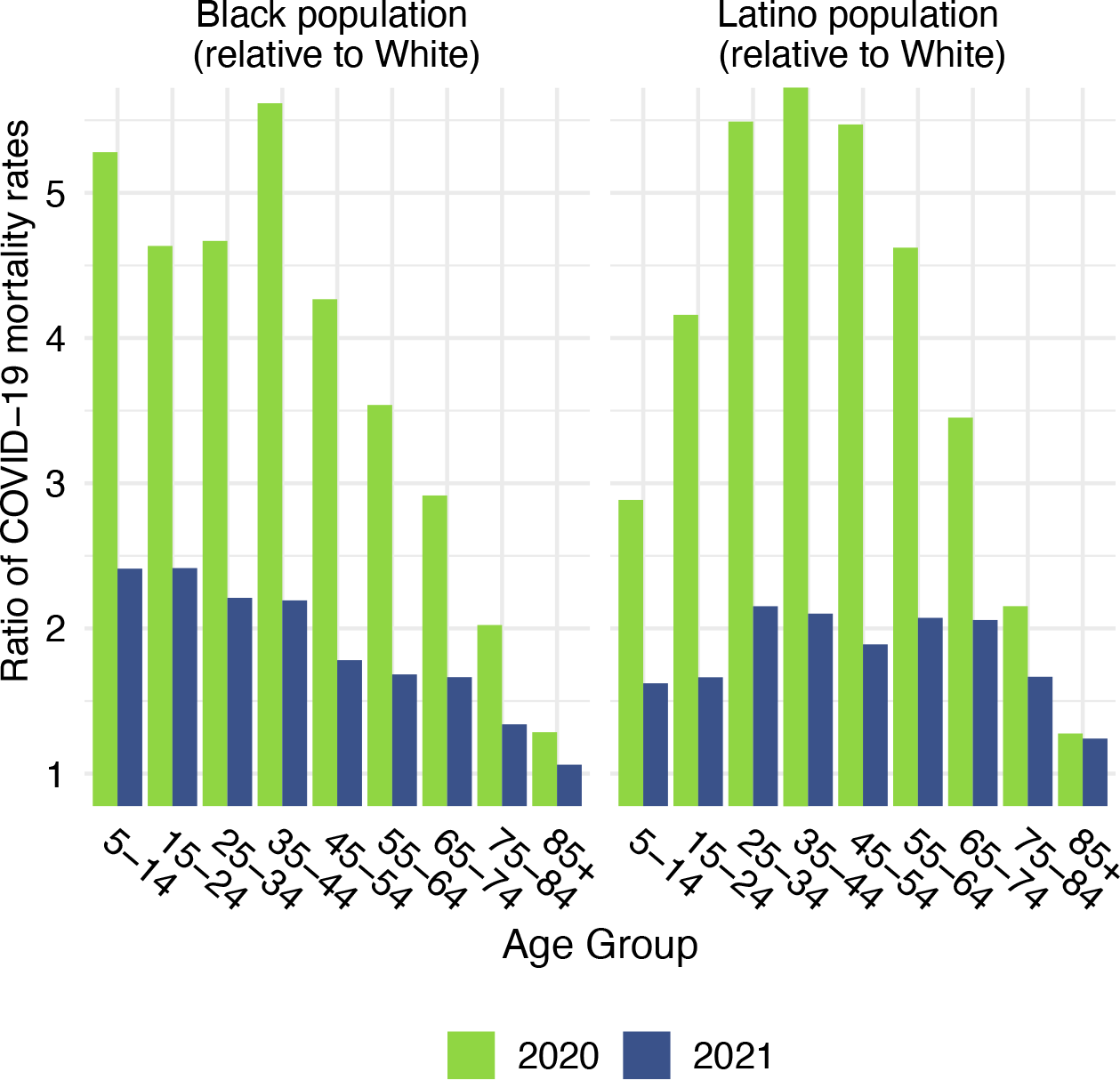
Ratio of age-specific COVID-19 mortality rates for the Black and Latino populations relative to the White population, 2020 and 2021. Data are from CDC WONDER (July 6, 2022, update). Ratios below age 5 are not shown because some counts are suppressed.

Life expectancy estimates for the total US population and by race/ethnicity are displayed in Table 1. For reference, official 2019 life expectancy values are displayed in panel A. Estimates of 2020 life expectancy are displayed in panel B and those for 2021 in panel C. To facilitate comparisons between 2020 and 2021 and highlight the persistent racial/ethnic disparities, Fig 4 displays the magnitudes of the reductions in life expectancy at birth and at age 65 for 2020 and 2021.

**Table 1:** Life expectancy estimates and reductions (in years) from 2019 for the total US population and by race/ethnicity.

|  | **Total Population** |  | **non-Latino White** |  | **non-Latino Black** |  | **Latino** |  |
| --- | --- | --- | --- | --- | --- | --- | --- | --- |
|  | Birth | Age 65 | Birth | Age 65 | Birth | Age 65 | Birth | Age 65 |
| **A) Pre-pandemic** |  |  |  |  |  |  |  |  |
| 2019 $e_x$ | 78.8 | 19.6 | 78.8 | 19.5 | 74.8 | 18.2 | 81.9 | 21.6 |
| **B) 2020 estimates** |  |  |  |  |  |  |  |  |
| Estimated 2020 $e_x$ | 77.2 | 18.8 | 77.5 | 18.7 | 71.7 | 16.5 | 78.7 | 19.7 |
| Change relative to 2019 $e_x$ | -1.6 | -0.8 | -1.3 | -0.8 | -3.1 | -1.7 | -3.2 | -1.9 |
| **C) 2021 estimates** |  |  |  |  |  |  |  |  |
| Estimated 2021 $e_x$ | 76.6 | 18.8 | 76.8 | 18.6 | 71.3 | 17.0 | 78.2 | 20.1 |
| Change relative to 2020 $e_x$ | -0.6 | 0 | -0.7 | -0.1 | -0.4 | 0.5 | -0.5 | 0.4 |
| Change relative to 2019 $e_x$ | -2.2 | -0.8 | -2.0 | -0.9 | -3.5 | -1.2 | -3.7 | -1.5 |

Notes: Apart from life expectancy ( $e_x$ ) values from 2019 that are provided by the National Vital Statistics System, all life expectancy estimates are authors' calculations. Estimates for 2021 are based on provisional mortality data provided by CDC WONDER.

**Fig 4.**
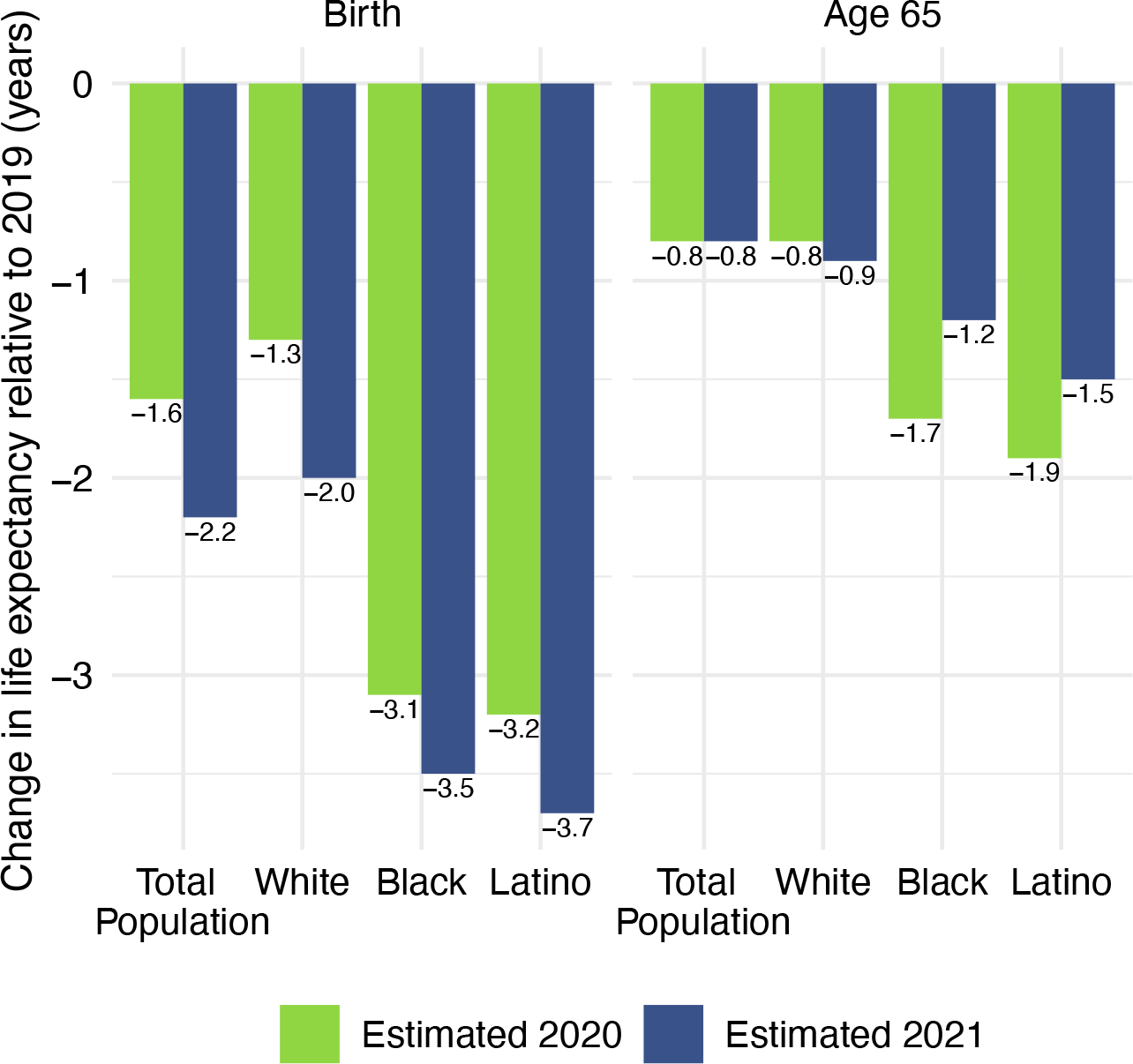
Reduction in life expectancy at birth and at age 65 (years) relative to 2019 by race/ethnicity, 2020 and 2021. Changes are all relative to 2019 life expectancy. Data are from CDC WONDER (July 6, 2022, update).

These estimates indicate a 2.2-year reduction in life expectancy at birth and a 0.8-year reduction in life expectancy at age 65 in 2021 for the total US population relative to 2019. The reductions in life expectancy at birth are largest for the Latino population (3.7 years), followed by the Black population (3.5 years), and smallest for the White population (2.0 years). The estimated 2021 reductions in life expectancy at birth for all populations exceed the reductions estimated for 2020, but the changes are largest for Whites (an additional 0.7-year decline between 2021 and 2021). The additional reductions for the Black and Latino populations are more moderate (0.4 and 0.5 years respectively), but far from negligible. Corresponding estimates for Native Americans presented elsewhere show an even more pronounced worsening of the mortality impact of COVID-19: a loss in life expectancy at birth that increased dramatically from 4.5 years in 2020 to 6.4 years in 2021 relative to 2019, i.e., an additional 1.9-year reduction, with losses in both years vastly exceeding those for the Black and Latino populations (22).

The estimated reductions in life expectancy at birth for the Black and Latino populations in 2021 relative to 2019 are 1.8 and 1.9 times, respectively, the 2-year reduction for Whites. In 2020, the reductions for both the Black and Latino populations were nearly 2.5 that of Whites. Although these estimates of loss in life expectancy relative to Whites in 2021 are not quite as severe as in 2020, they reveal another year of pronounced racial/ethnic inequities underlying an even more severe overall impact of the COVID-19 pandemic on life expectancy.

## Discussion

With the advent of highly effective vaccines to protect against COVID-19, many in the scientific and public health communities had hoped that the mortality impact of COVID-19 would be substantially lessened in 2021 with period life expectancy converging toward pre-pandemic levels. However, our findings reveal a devasting mortality impact of the pandemic in 2021, one that exceeds that in 2020 for all populations, with persistently large racial and ethnic disparities.

Our estimates in Table 1 reveal no difference between the decline in life expectancy at age 65 estimated for the total population in 2021 and that estimated for 2020 (both relative to 2019) – the slightly larger impact for Whites in 2021 is counteracted by improvements among the Black and Latino populations. This pattern contrasts with the decline in life expectancy at birth in 2021 and is consistent with the shifting distribution of COVID-19 deaths toward younger ages in 2021 (Fig 1). The younger age distribution of COVID-19 deaths is largely a consequence of the steady increase in the prevalence of vaccination by age, with high levels achieved for the elderly (over 85% of the 65 and over population fully vaccinated by the end of 2021), in contrast to lower coverage among younger adults (approximately 63% of adults aged 25-39 fully vaccinated by the end of 2021) (4,7). High vaccination rates among nursing home residents, who are particularly vulnerable to adverse COVID-19 outcomes, paired with stricter infection protocols, also helped reduce the mortality impact of COVID-19 on older adults in 2021 (23,24). The net result, as shown in Fig 2, is that mortality from COVID-19 for each racial/ethnic group declined substantially at the oldest ages but changed relatively little in young and middle age groups. Although differences between 2020 and 2021 in age-specific death rates in young and middle age groups appear small in Fig 2, the modest increases throughout much of this age range, most notable for Whites, likely make a relatively large contribution to the change in life expectancy at birth between 2020 and 2021.

These results underscore the persistence of large racial/ethnic disparities in the effect of COVID-19 on life expectancy. The disproportionately high losses of life in the Black and Latino populations reflect the social and economic inequities that have been repeatedly acknowledged throughout the pandemic, most notably high rates of poverty and crowded housing, low income jobs that cannot be performed remotely, a high prevalence of chronic health conditions, and inadequate access to quality healthcare (8,9,11). Despite having had higher life expectancy than Whites prior to the pandemic, Latinos suffered huge losses in life expectancy in both 2020 and 2021. High COVID-19 death rates for this population have been partly attributed to relatively low levels of health insurance coverage, higher prevalence of multigenerational households than most other groups, and language barriers to obtaining information on viral transmission and mitigation strategies (9,10,12,25). In addition, Latino workers suffered disproportionate job and income losses during the pandemic because of their overrepresentation in the gig economy and in industries greatly impacted during this period (e.g., construction and leisure and hospitality) and because many Latinos were ineligible for government benefits (26). Although data on race and ethnicity of vaccine recipients are incomplete, existing information suggests that the persistent racial/ethnic disparities are likely partially the result of differences in vaccine uptake early in 2021. For example, after taking differences in age structure into account, Reitsma and colleagues estimate that vaccine uptake rates (for at least one dose) were about 30% higher in Whites than in the Black and Latino populations through the end of March, 2021, with huge variability across states (27).

It is important to emphasize that, although the estimates of life expectancy reductions in Table 1 indicate a slight narrowing of the racial/ethnic differentials from the previous year, this is largely due to greater absolute and relative life expectancy reductions in the White population compared with the corresponding reductions in the Black and Latino populations. The worsening life expectancy in 2021 among Whites is likely largely attributable to higher COVID-19 mortality in 2021, which could reflect lower adherence to social distancing and masking guidelines relative to other races/ethnicities (28–30). Although all groups reported high adherence to public health guidelines at the beginning of the pandemic, Whites appear to have resumed social activities and ceased mask-wearing more quickly than Black and Latino individuals (28,29).

Although COVID-19 is the primary cause of continued life expectancy reductions in 2021, increases in other causes of death relative to pre-pandemic levels contribute to these life expectancy declines. In particular, the pandemic has been cited as a source of much of the upsurge in drug overdose deaths in 2020 and 2021, exacerbating the increase in drug overdose that began during the years prior to the pandemic (31). Greater availability and use of synthetic opioids in recent years, especially illicitly manufactured fentanyl and fentanyl analogs which are often mixed or used together with other substances and are especially lethal, have led to a greatly enhanced risk of drug overdose (32,33). Increased mortality rates from other non-COVID-19 causes, including heart disease and diabetes, may have resulted from increased severity of co-morbidities due to COVID-19 infection, delays in primary and preventive care or reduced disease management, and inadequate healthcare due to shortages of equipment, staff, and space, as occurred in 2020 (34–37). In 2020, increases in these causes of death exceeded decreases in other causes (e.g., cancer, Alzheimer’s disease, chronic lower respiratory diseases), contributing to decreases in life expectancy beyond those implied by COVID-19 deaths alone (1,2,18,31).

This analysis is subject to several limitations. Our estimates for 2021 life expectancy rely on provisional mortality data, which are subject to reporting and processing delays, although NVSS estimates that mortality data are over 99% complete after three months (1). Additionally, the publicly available data do not allow us to disaggregate mortality rates at age 85 and above into narrower age intervals and do not incorporate adjustments for age misreporting or underreporting of mortality at older ages, which leads to underestimates mortality rates at these older ages. Because the published estimates for 2019 incorporate these adjustments, the resulting overestimates of life expectancy for 2020 and 2021 lead to underestimates of the corresponding losses during the pandemic.

As period measures, our estimates of life expectancy reductions for 2021 quantify loss of life resulting from the pandemic along with disparities across racial and ethnic groups, but it is important to recognize that they do not represent expectations of remaining life for any living cohort, which depend on future mortality conditions. It is uncertain whether life expectancy in 2022 will show a substantial improvement from the past two years, but there are several reasons for optimism. The number of deaths from COVID-19 in the first six months of 2022 is approximately 30% lower than during the corresponding months of 2021 (38). The administration of additional booster doses should help protect many at high-risk of dying from complications of COVID-19 (39,40). Other efforts that should reduce the mortality impact of COVID-19 are the ongoing development of different types of vaccines, including those targeting new and multiple variants, the expansion of vaccine eligibility to young children, and the increasing availability of effective oral self-administered antiviral treatments, which can reduce the risk that a COVID-19 infection develops into severe disease (41). Many high-income countries that had substantial losses in life expectancy at birth in 2020 experienced less severe reductions in 2021, and some, including France, Belgium, Switzerland, and Sweden, appear to have returned to pre-pandemic life expectancy in 2021 (42).

However, there are also many reasons why 2022 may see continued elevated mortality levels and persistent inequities in the US. At the time of writing in July 2022, all states have ended indoor mask mandates as part of a strategy of “living with COVID” or treating it as endemic (43). Such a strategy may entail continued high COVID-19 mortality for several reasons. There is still substantial vaccine refusal in the US, with approximately 15% of adults not having received any dose of a COVID-19 vaccine by the end of 2021 (4), and vaccine refusal is unlikely to be substantially diminished in the near future. The appearance of a highly transmissible Omicron variant at the end of 2021 resulted in another large surge of deaths, despite having lower fatality than previous variants, and even more transmissible subvariants of Omicron continue to emerge (4,6). There is also a constant threat of new variants that, as with Omicron, will be at least partly resistant to existing vaccines and perhaps existing treatments, and that may be more fatal.

Apart from direct COVID-19 deaths, there is also evidence that survivors of COVID-19 have increased mortality risks for at least six months following initial recovery (44), and the mortality impact of long COVID is not yet known. In addition, increased risks of dying from a broad range of conditions may have been triggered by detrimental changes in health-related behaviors induced by the many social and economic stressors during the pandemic; these behaviors include higher rates of smoking, drinking and drug use; worse nutrition; and reduced exercise (45–47). And, unfortunately, other long-term impacts of the pandemic on mortality resulting from the many social, economic, and healthcare disruptions during the past two years will likely continue to disproportionately affect vulnerable populations.

## Data Availability

The data used in this study are all publicly available through the provided links.

https://wonder.cdc.gov/mcd-icd10-provisional.html

